# Validation of dementia care-related scales among informal caregivers of Latinos with dementia or mild cognitive impairment

**DOI:** 10.1101/2024.08.28.24312743

**Authors:** Jaime Perales-Puchalt, Irene Checa, Begoña Espejo, Marta de la C. Martín Carbonell, Mónica Fracachán-Cabrera, Christina Baker, Mariana Ramírez-Mantilla, Prisca Mendez-Asaro, Malissia Zimmer, Kristine Williams, K. Allen Greiner, Jana Zaudke, Hector Arreaza, Idaly Velez-Uribe, Henry Moore, Vanessa Sepulveda-Rivera, Kylie Meyer, Donna Benton, Krystal Kittle, Lindsey Gillen, Jeffrey M Burns

## Abstract

**Objectives:** To test the psychometric properties of several dementia care-related scales among Latinos in the US.

**Design:** We leveraged secondary baseline data from a one-arm mHealth trial on dementia caregiver support. We included 100 responses for caregiver-focused scales and 88 responses for care recipient-focused scales. Scales included the Neuropsychiatric Inventory Questionnaire Severity and Distress scales, six-item Zarit Burden Inventory, Ten-item Center for Epidemiologic Studies Depression Scale, Geriatric Depression Inventory, Quality of Life in Alzheimer’s Disease, and Single-item Satisfaction With Life Scale. We calculated concurrent validity using Pearson and Spearman correlations and expected correlations amongst all variables in line with the Stress Process Framework. We calculated internal consistency reliability using Cronbach’s alpha.

**Results:** All concurrent validity correlations followed the expected directionality, with 19/21 inter-scale correlations in the total sample reaching statistical significance (p<0.05), and 17/21 reaching at least a low correlation (0.3). Cronbach’s alpha ranged from 0.832 to 0.879 in all scales in the total sample.

**Conclusion:** The English and Spanish caregiver-administered scales tested in this manuscript have good psychometric properties.

**Clinical Implications:** The dementia care-related scales are now appropriately available for use among US Latinos in research and clinical contexts.

## Introduction

Eliminating dementia disparities among Latinos is crucial. Dementia is a major cause of mortality and disability in later life, and costs the US healthcare system more than cancer or heart disease.^1,2^ Dementia also impacts family caregivers, to the extent that ∼40% of have depressive or anxiety disorders.^3,4^ The National Alzheimer’s Plan Act and the National Institutes of Health have identified addressing dementia disparities among ethnic minorities as a public health priority.^5^ Latinos are the largest minoritized ethnoracial group and the fastest growing regarding older age in the US. ^6–8^ Despite their large representation in the US, Latinos experience multiple disparities in dementia, including a disproportionately high dementia risk,^9^ low and delayed detection,^10^ and poorer treatment and care.^11–13^ These disparities are compounded by a scarcity of research that could shed light on how to eliminate these disparities.^14,15^

A factor that can contribute to poorer care and a lack of research includes not having the appropriate scales to measure dementia care outcomes among Latinos. Every racial and ethnic group possesses distinctive cultural traits, encompassing values, norms, and attitudes.^16,17^ For this reason, it is important to determine if instruments that were originally developed with primarily non-Latino White populations in the US are accurate and valid among Latinos.^18^ In addition to cultural aspects, in the US, 73% of Latinos report speaking Spanish at home, and 47% of Latino older adults report having poor English proficiency. Despite these facts, the practice of applying standard measures among Latinos without exploring their psychometric properties continues to be common practice.^18^

Some constructs that are key for dementia care include depressive symptomatology, caregiver burden, neuropsychiatric symptom severity and distress, quality of life, and life satisfaction.^19^ Validated scales exist for some of these constructs for US Latinos in the general or non-dementia population. These scales include the Center for Epidemiologic Studies Depression Scale 10 (CES-D 10),^20^ and the Patient Health Questionnaires (PHQ) 9 and 2 for depression,^21^ or the Problem Areas In Diabetes to emotional distress.^22^ However, most of these domains have not been validated in the dementia care context.^23^ Validating short versions of existing scales (e.g., Zarit Burden Inventory; ZBI 6),^24,25^ is important to ease their implementation into clinical and research settings, reduce clinic time constraints, and reduce patient/participant burden. In this manuscript, we aim to test the psychometric properties of several dementia care-related scales among Latinos. These scales are relevant in dementia care research and have been used in several intervention and observational studies.^19,26–29^

## Methods

The current secondary analysis uses baseline data from a one-arm pre-post-intervention trial design. The original protocol included only participants referred by partner clinics. However, due to the slow pace of referrals, the research team adapted the protocol to allow recruitment from any source. We enrolled caregiver-care recipient dyad participants from May 2022 to February 2024 from our center’s clinic (6.0%), previous research studies (24.0%), partnering clinic referrals (23.0%), partner community institution referrals (12.0%), direct community outreach via “Promotoras de Salud” or health events (12.0%), research colleague referrals (8.0%), web registries and ads (13.0%), and word of mouth (2.0%). Participants were eligible if both caregiver and care recipient spoke Spanish or English. Caregivers were eligible if they were able to understand the informed consent via seven yes/no questions, were 18 or older and were a relative or friend of the care recipient for whom they provided some sort of care or support.

Additional caregiver inclusion criteria included contact with the care recipient at least once a week in-person or via phone, and ownership of a cellphone with a flat fee for Short Message Service (SMS) text messaging. Care recipients were eligible if they or their caregiver identified them as Latino, had a clinical or research dementia/mild cognitive impairment diagnosis, and attended a primary care clinic. In our previous research, advisory board members suggested that if two or more people cared for a single person with dementia, they were included in the study, as this approach could reduce burden and increase social support ^30^. For this reason, we allowed more than one dyad per person with dementia. All study procedures were approved by the Institutional Review Board of the University of Kansas Medical Center (STUDY00145615). All caregiver participants gave written informed consent, as did those care recipients who were determined to be able to respond on their own via questions on comprehension of the informed consent. This study was registered in ClinicalTrials.gov under the ID NCT04418232.

### Procedures

The research team provided an overview of the study’s key aspects to potential participants through phone discussions or secure video calls. Those expressing interest underwent a screening process to determine eligibility. Once deemed eligible, caregiver participants (and individuals with dementia, if they could independently consent according to the caregiver and consent form-related inquiries) were invited to electronically sign an informed consent form and arrange a follow-up phone or video session to complete the initial assessment. Caregivers supplied all necessary information about themselves and acted as proxies for people with dementia who were unable to provide consent. Upon completion of the baseline assessments, all participants were officially enrolled in the study and promptly commenced the Alianza Latina intervention, with a 6-month follow-up survey scheduled.

### Intervention

Alianza Latina is a six-month bilingual intervention, designed to cater to the unique needs of Latino caregivers. It integrates *CuidaTEXT*, an SMS program, with monthly phone consultations facilitated by a trained coach from the research team. These consultations aim to identify unaddressed needs and provide necessary support. Participants in *CuidaTEXT* receive scheduled messages and can also request on-demand assistance by texting. The intervention, and its development process, have been extensively detailed previously. ^30^ *CuidaTEXT* entails sending 1-3 automated daily messages covering various aspects such as logistics, dementia education, self-care, social support, end-of-life care, managing dementia-related behaviors, and problem-solving strategies. Additionally, participants can text keyword-based queries for immediate assistance on the above-mentioned topics and engage in live chat sessions with the coach for further guidance upon request.

### Assessment

The data discussed in this paper solely pertains to the baseline assessments. Socio-demographic, acculturation and relation information included the caregivers’ years of age, gender, ethnicity, race, country of birth, years living in the US if born in a foreign country, US region of residence (e.g., Midwest), years of education, medical insurance status, medical care status, primary language (e.g., Spanish, English, both), English proficiency (5 point Likert scale ranging from very low to very high), marital status, working status, difficulty to pay for basic needs (5 point Likert scale ranging from very easy to very difficult), and relation to the care recipient. Most of this information was also gathered for the person with dementia.

We tested the psychometric properties of the following scales in their English and Spanish language versions:

Neuropsychiatric Inventory Questionnaire severity (NPI-Q-S; care recipient) and distress (NPI-Q-D; caregiver). The NPI-Q is a clinical instrument for evaluating psychopathology in dementia with two scales, care recipient severity (NPI-Q-S) and caregiver distress (NPI-Q-D). If any of the 12 neuropsychiatric symptoms are present in the last month (e.g., depression, repeating), caregivers rate the level of severity for the IWDs on a 3-point scale (Mild–Severe). An overall severity summary score is calculated by adding the severity scores of all items. For any present symptom, caregivers also rate their own distress on a 6-point scale (Not Distressing at All- Extreme or Very Severe Distress). An overall distress summary score is calculated by adding the distress scores of all items. Higher scores indicate higher severity and distress. In the current study, we used the original NPI-Q version in English,^31^ and combined the NPI-Q-S from an adapted US Spanish version from the National Alzheimer’s Coordinating Center,^32^ and the NPI-Q-D from the adapted Spanish version used in different Latin American countries (Cuba, Dominican Republic, Peru, Mexico, Venezuela, and Puerto Rico) for the 10/66 Study.^33^

Six-item Zarit Burden Inventory (ZBI-6; caregiver). The ZBI measures caregiver burden (e.g., having enough time to yourself). Originally designed and tested in 1980 in English in a US sample containing 29 items, it was later reduced to six items and tested in English in the UK, without modifying the language in the items.^34^ Each of the six items of the ZBI-6 is a statement the caregiver is asked to endorse using a 5-point scale. Response options range from 0 (Never) to 4 (Nearly Always). An overall burden summary score is calculated by adding the scores of all items, and higher scores indicate a higher burden. We used the UK 6-item version in English,^34^ and extracted the same items in Spanish from a translation from Spain that we considered appropriate for US Spanish speakers.^35^

Ten-item Center for Epidemiologic Studies Depression Scale (CES-D 10; caregiver). The CES-D is a scale that measures depressive symptomatology. Originally designed and tested in 1977 in English in a US sample with 20 items,^36^ which were later reduced to 10 items,^37^ and tested in Chinese.^38,39^ This reduced scale was later adapted and tested in English and Spanish among a general US Latino adult population sample in the Study of Latinos.^20^ The CES-D-10 is a 10-item, self-report rating scale that measures characteristic symptoms of depression in the past week (e.g., depression, loneliness). Each item is rated on a 4-point scale, from 0 (Rarely or None of the Time) to 3 (Most or All of the Time) with positively worded items (items 5 and 8) reverse scored. Items yield summary scores that range from 0 to 30, with higher scores indicating higher depression severity. We used the CES-D-10 in English and Spanish from the Study of Latinos for the current study.^20^

Single-item Satisfaction With Life Scale (SWLS). The SWLS was developed in 2014 and validated in three representative samples in the US and Germany.^40^ The item asks “In general, how satisfied are you with your life?” and is rated on a 4-point scale, from 1 (very dissatisfied) to 4 (very satisfied). Both English and Spanish versions were extracted from the 2010 Behavioral Risk Factor Surveillance System.^41^

Geriatric Depression Scale (GDS; care recipient). The GDS is a scale that measures depressive symptomatology tailored to older adults. Originally designed and tested in 1982 in English in a US sample containing 30 items,^42^ it was later reduced to 15 in 1986 in the same population.^43^ Participants are asked to respond to each item by answering yes or no about how they felt over the past week. Of the 15 items, 10 indicate the presence of depression when answered positively, while the rest (question numbers 1, 5, 7, 11, 13) indicate depression when answered negatively. An overall summary score is calculated by adding the scores of all items, and higher scores indicate higher depressive symptomatology. In the current study, we used the original 1986 version in English and an adapted US Spanish version from the National Alzheimer’s Coordinating Center.^32,43^

Quality of Life in Alzheimer’s Disease (QOL-AD; care recipient). The QOL-AD measures health-related quality of life tailored to Alzheimer’s disease. The scale was designed and tested in 2002 in English in a US sample and contains 13 items.^44^ This scale uses a scale of 1–4 (poor, fair, good, or excellent) to rate a variety of life domains, including the patient’s physical health, mood, relationships, activities, and ability to complete tasks. An overall summary score is calculated by adding the scores of all items, and higher scores indicate higher health-related quality of life. We used the original USA English version,^44^ and a Spanish version from Mexico that we considered to be appropriate for US Spanish speakers.^45^

### Analysis

For the current analysis, we included data reported by caregivers on their behalf or data reported by caregivers on their care recipients’ behalf. We excluded Care recipients’ self-reported data because very few data (n=12) were completed by care recipients and research shows self and proxy-ratings might measure different constructs.^46^ Therefore, we included 100 responses for caregiver-focused scales and 88 responses for care recipient-focused scales. We performed analyses using IBM Statistical Package for the Social Sciences (SPSS) Version 22.^47^ We calculated means and standard deviations, or frequencies and percentages for descriptive statistics. We calculated concurrent validity using Pearson and Spearman correlations, based on their normal or non-parametric distribution, and used a significance level of α= 0.05 to protect against type I error. We consider correlations to be low if they range from 0.3 to 0.5; moderate if they range from 0.5 to 0.7; and high if they are higher than 0.7.^48^ We expect these constructs to correlate amongst themselves in line with the Stress Process Framework.^19,49^ This framework poses that caregivers’ stressors (e.g., behavioral symptoms of the care recipient) have health consequences (e.g., caregiver depression). We calculated internal consistency reliability using Cronbach’s alpha, which values include ≤0.5 (unacceptable), 0.7 (acceptable), 0.8 (good), and ≥0.9 (excellent).^50^ We report analyses in the total sample and stratify them by the language used by the caregiver to complete the scales (English or Spanish).

## Results

Table 1 shows the characteristics of the total sample, stratified by the language in which participants responded to the assessments. Caregivers were on average 52.0 years old (standard deviation [SD] 10.2). Ninety-five caregivers identified as Latino (95.0%), 82 as women (82.0%), 43 were born in Mexico (43.0%), and 30 were born in a different Latin-American country (30.0%). Fifty-five caregivers spoke Spanish as their primary language (55.0%), 72 were married or lived with a partner (72.0%), and 31 found it difficult or very difficult to pay for basic needs (31.0%). Seventy-three caregivers identified as the adult children of the person with dementia (73.0%). Caregivers were on average 78.4 years old (SD 9.9). All identified as Latino, 64 as women (72.7%), 47 were born in Mexico (53.4%), and 33 were born in a different Latin-American country (37.5%). Seventy-seven caregivers spoke Spanish as their primary language (87.5%), and 24 were married or lived with a partner (27.3%). The distribution of characteristics was similar except for caregiver and care recipient country of birth, primary language, years in the US, educational achievement, and medical insurance, caregivers’ region of residence, and care recipients’ medical care access.

**Table 1.** Baseline characteristics of the participants enrolled in *Alianza Latina*.

| Caregivers | Total<br>(n=100) | Spanish<br>speakers<br>(n=64) | English<br>Speakers<br>(n=36) |
| --- | --- | --- | --- |
| Age in years, mean (SD) | 52.0 (10.2) | 50.9 (8.7) | 54.1 (12.2) |
| Women, % (n) | 82.0% (82) | 84.4% (54) | 77.8% (28) |
| Latino ethnicity, % (n) | 95.0% (95) | 96.9% (62) | 91.7% (33) |
| Race, % (n) |  |  |  |
| Other, % (n) | 71.0% (71) | 76.6% (49) | 61.1% (22) |
| White, % (n) | 25.0% (25) | 20.3% (13) | 33.3% (12) |
| Multiple races, % (n) | 2.0% (2) | 1.6% (1) | 2.8% (1) |
| Black, % (n) | 1.0% (1) | 0.0% (0) | 2.8% (1) |
| Don't know, % (n) | 1.0% (1) | 1.6% (1) | 0.0% (0) |
| Country of birth |  |  |  |
| US, % (n) | 27.0% (27) | 10.9% (7) | 55.6% (20) |
| Mexico, % (n) | 43.0% (43) | 56.3% (36) | 19.4% (7) |
| Other, % (n) | 30.0% (30) | 32.8% (21) | 25.0% (9) |
| Years in the US among those born abroad, m (SD) | 28.8 (14.2) | 25.8 (11.1) | 39.8 (18.7) |
| USA region or residence |  |  |  |
| Midwest, % (n) | 70.0% (70) | 81.3% (52) | 50.0% (18) |
| West, % (n) | 21.0% (21) | 14.1% (9) | 33.3% (12) |
| South, % (n) | 6.0% (6) | 3.1% (2) | 11.1% (4) |
| East, % (n) | 3.0% (3) | 1.6% (1) | 5.6% (2) |
| Years of education, m (SD) | 12.6 (4.4) | 11.3 (4.7) | 14.8 (2.6) |
| Without medical insurance, % (n) | 34.0% (34) | 45.3% (29) | 13.9% (5) |
| No regular source of medical care, % (n) | 17.0% (17) | 15.6% (10) | 19.4% (7) |
| Spanish only as primary language, % (n) | 55.0% (55) | 81.3% (52) | 8.3% (3) |
| Married or have a partner, % (n) | 72.0% (72) | 71.9% (46) | 72.2% (26) |
| Currently working, % (n) | 64.0% (64) | 64.1% (41) | 63.9% (23) |
| Difficult or very difficult to pay for basic needs, % (n) | 31.0% (31) | 32.8% (21) | 27.8% (10) |
| Relation to care recipient |  |  |  |
| Children, % (n) | 73.0% (73) | 76.6% (49) | 66.7% (24) |
| Partner, % (n) | 11.0% (11) | 7.8% (5) | 16.7% (6) |
| Children-in-law, % (n) | 4.0% (4) | 4.7% (3) | 2.8% (1) |
| Other, % (n) | 12.0% (12) | 11.0% (7) | 13.9% (5) |
| Care recipients (with a proxy respondent) | Total (n=88) | Spanish speakers (n=57) | English Speakers (n=31) |
| Age in years, m (SD) | 78.4 (9.9) | 78.9 (8.3) | 77.3 (12.5) |
| Woman, % (n) | 72.7% (64) | 77.2% (44) | 64.5% (20) |
| Latino ethnicity, % (n) | 100.0% (88) | 100.0% (57) | 100.0% (31) |
| Race, % (n) |  |  |  |
| Other, % (n) | 80.7% (71) | 80.7% (46) | 80.6% (25) |
| White, % (n) | 18.2% (16) | 19.3% (11) | 16.1% (5) |
| Black, % (n) | 1.1% (1) | 0.0% (0) | 3.2% (1) |
| Country of birth |  |  |  |
| US, % (n) | 9.1% (8) | 0.0% (0) | 25.8% (8) |
| Mexico, % (n) | 53.4% (47) | 59.6% (34) | 41.9% (13) |
| Other, % (n) | 37.5% (33) | 40.4% (23) | 32.3% (10) |
| Years in the US among those born abroad, m (SD) | 35.5 (20.5) | 30.7 (19.6) | 47.6 (18.0) |
| Years of education, m (SD) | 14.0 (25.6) | 12.1 (24.5) | 17.5 (27.5) |
| Without medical insurance, % (n) | 8.0% (7) | 12.3% (7) | 0.0% (0) |
| No regular source of medical care, % (n) | 9.1% (8) | 12.3% (7) | 3.2% (1) |
| Spanish only as primary language, % (n) | 87.5% (77) | 100.0% (57) | 64.5% (20) |
| Married or have a partner, % (n) | 27.3% (24) | 22.8% (13) | 35.5% (11) |
| Currently working, % (n) | 1.1% (1) | 0.0% (0) | 3.2% (1) |

Table 2 shows the concurrent validity of all scales, as calculated by Pearson and Spearman correlations. All correlations followed the expected directionality and out of the 21 inter-scale correlations conducted in the total sample, 19 were statistically significant (p<0.05), 17 reached at least the 0.3 level (low), six the 0.5 level (moderate), and one the 0.7 level (high). Correlations followed similar patterns in the groups that responded in Spanish and English.

Table 3 shows the internal consistency of all scales except for the single-item SWLS.

Cronbach’s alpha ranged from 0.832 to 0.879 in all scales in the total sample, indicating good internal consistency. Patterns remained similar for scales completed in Spanish and English, although alphas decreased or increased slightly in some cases (e.g., 0.895 for the Spanish NPI-Q-D or 0.763 for the English ZBI-6).

**Table 2.** Concurrent validity correlations among scales.

| All languages | Single-item SWLS <sup>a</sup> |  |  | NPI-Q S <sup>b</sup> |  |  | NPI-Q D <sup>a</sup> |  |  | ZBI-6 <sup>a</sup> |  |  | CES-D 10 <sup>a</sup> |  |  | GDS <sup>b</sup> |  |  | QOL-AD <sup>b</sup> |  |  |
| --- | --- | --- | --- | --- | --- | --- | --- | --- | --- | --- | --- | --- | --- | --- | --- | --- | --- | --- | --- | --- | --- |
|  | Total | Spanish | English | Total | Spanish | English | Total | Spanish | English | Total | Spanish | English | Total | Spanish | English | Total | Spanish | English | Total | Spanish | English |
| Single-item SWLS <sup>a</sup> | 1 | 1 | 1 | - | - | - | - | - | - | - | - | - | - | - | - | - | - | - | - | - | - |
| NPI-Q S <sup>b</sup> | -0.276** | -0.288* | -0.285 | 1 | 1 | 1 | - | - | - | - | - | - | - | - | - | - | - | - | - | - | - |
| NPI-Q D <sup>a</sup> | -0.326** | -0.355** | -0.165 | 0.729** | 0.749** | 0.747** | 1 | 1 | 1 | - | - | - | - | - | - | - | - | - | - | - | - |
| ZBI-6 <sup>a</sup> | -0.485** | -0.538** | -0.332* | 0.420** | 0.500** | 0.394* | 0.592** | 0.636** | 0.374* | 1 | 1 | 1 | - | - | - | - | - | - | - | - | - |
| CES-D 10 <sup>a</sup> | -0.577** | -0.514** | -0.655** | 0.361** | 0.408** | 0.169 | 0.524** | 0.521** | 0.339* | 0.694** | 0.706** | 0.587** | 1 | 1 | 1 | - | - | - | - | - | - |
| GDS <sup>b</sup> | -0.606** | -0.542** | -0.713** | 0.394** | 0.406** | 0.468** | 0.429** | 0.453** | 0.530** | 0.200 | 0.334* | 0.084 | 0.156 | 0.252 | 0.045 | 1 | 1 | 1 | - | - | - |
| QOL-AD <sup>b</sup> | 0.453** | 0.405** | 0.531** | -0.379** | -0.382** | -0.440* | -0.386** | -0.416** | -0.502** | -0.250* | -0.324* | -0.337 | -0.369** | -0.415** | -0.420* | -0.681** | -0.693** | -0.668** | 1 | 1 | 1 |

a: rating by caregiver about themselves; b: rating by caregivers about their care recipient. Pearson correlations used for all variables except for NPI-Q S and D, for which we used Spearman correlations. \* Correlation is significant at the 0.05 level; \*\* Correlation is significant at the 0.01 level.

**Table 3.** Internal consistency reliability of scales.

|  | NPI-Q S <sup>a</sup> | NPI-Q D <sup>b</sup> | ZBI-6 <sup>a</sup> | CES-D 10 <sup>a</sup> | GDS <sup>b</sup> | QOL-AD <sup>b</sup> |
| --- | --- | --- | --- | --- | --- | --- |
| Total sample | 0.834 | 0.879 | 0.876 | 0.832 | 0.834 | 0.834 |
| Spanish | 0.857 | 0.895 | 0.798 | 0.830 | 0.807 | 0.811 |
| English | 0.784 | 0.828 | 0.763 | 0.808 | 0.878 | 0.865 |

a: rating by caregiver about themselves; b: rating by caregivers about their care recipient.

## Discussion

To our knowledge, this is the first study to explore the psychometric properties of the English and Spanish versions of the ZBI-6, NPI-Q, SWLS, QOL-Ad and GDS in a US Latino population. This is also the first study to explore the psychometric properties of all the included scales in a Latino sample in the context of dementia.^23^ We used baseline data from a support trial for family caregivers of Latinos with dementia to test the scales’ reliability and validity.

Based on the Stress Process Framework, we hypothesized as part of the concurrent validity assessment that the scales would correlate amongst themselves.^19^ Most correlations supported our concurrent validity hypothesis, and the internal consistency was good for all scales.

Consistent with previous studies with non-Latino populations, the scales had equivalent internal consistency reliability.^39^ For example, the CES-D-10 achieved an internal consistency of 0.78 for the CES-D-10 in a community and psychogeriatric assessment clinic in China compared to 0.83 in our sample,^39^ the 15-item GDS achieved an internal consistency of 0.94 in two groups with and without a depression diagnosis in Greece compared to 0.834 in our sample,^51^ the NPI-Q-S achieved an internal consistency of 0.67 and the NPI-Q-D a 0.81 in a dementia clinic sample in Brazil compared with 0.83 and 0.88 respectively in our sample,^52^ the QOL-AD achieved an internal consistency of 0.82 in a sample with dementia in Mexico for the caregiver-administered version compared with 0.83 in our sample, and the ZBI-6 achieved an internal constancy of 0.83 in a sample with dementia in the UK compared with 0.88 in our sample.^34^

Most of the scales included in this manuscript have not previously been validated among Latinos in English or Spanish. The internal consistency reliability of the only scale we are aware of that was validated among Latinos, the CES-D-10, also aligns with the one found in Gonzalez et al,^20^ in which alpha was 0.82 among a general adult population sample of English and Spanish speaking Latinos in the US.^51^ Regarding the concurrent validity, most scales were correlated among each other. These correlations are in line with the relationships depicted in the Stress Process Framework,^49^ and previously tested in the Resources for Enhancing Alzheimer’s Caregiver Health intervention using structural equation modeling.^19^

This study has limitations. The sample size was based on a feasibility trial and therefore was not powered to validate these scales. However, the sample size is appropriate for calculating internal consistency in this manuscript with a minimum acceptable Cronbach’s alpha of 0.65 and an expected Cronbach’s alpha of 0.8;^53,54^ as well as to detect Pearson correlations of 0.28 or higher.^54–56^ Our limited sample size also impeded conducting confirmatory factor analyses or measurement invariance analyses. Analyzing the scales’ factor structure and measurement invariance is important to understand how different groups of scale items are related amongst them and to ensure that the measurement is equivalent among different groups who might differ in social desirability, interpretation of language, differential responses to items or other parameters.^57,58^ Given the nature of the study, we also did not assess test-retest reliability, which could have offered information with on the temporal accuracy of the measurement of the scales included. The sample is not probabilistic. Findings might not be generalizable to the US Latino caregiver population.

This study has implications for public health as well as research. Future epidemiological surveillance studies and trials can now use the data reported in this validation study to inform the scales they plan to select. The current study will be especially useful for studies that plan to assess health disparities among families with dementia that include US Latinos. Clinicians now have several validated tools to administer to caregivers of their Latino patients to assess relevant domains, including their patients’ quality of life, depression, and neuropsychiatric symptom severity, and their caregivers’ burden, depression, satisfaction with life and distress related to neuropsychiatric symptoms. These scales are relatively short, which makes them ideal in clinical settings that experience time constraints. Some ideas for future studies also stem from our findings. Future studies should test the factorial structure of the scales included in this manuscript among US Latinos. These studies should also analyze the measurement invariance by language and Latino descent (e.g., Mexican, Cuban, etc.). Studies such as the National Alzheimer’s Coordinating Center Unified Data Set might allow these analyses for two of the used scales (GDS and NPI-Q-S), given the larger number of Latino participants.^32^ Future studies should test the psychometric properties of the scales from this paper in a population sample such as the Study of Latinos (SOL), to increase the generalizability of findings.^20^

## Conclusions

The coming tsunami of Latinos with dementia will need their health and health related domains measured validly and reliably. The urgency of this need impacts both research, for which Latinos are under-represented, and clinical practice, where a likely biased one-size-fits-all approach is dominant. The current findings show that the English and Spanish caregiver-administered versions of the ZBI-6, NPI-Q-D, SWLS, and CES-D-10 for caregivers of Latinos with dementia and the caregiver-administered versions of the NPI-Q-D, QOL-AD, and GDS for Latino care recipients with dementia have good psychometric properties, namely internal consistency, and concurrent validity. These tools are now appropriately available for use among US Latinos in research and clinical contexts.

## Clinical Implications

- The English and Spanish caregiver-administered versions of the ZBI-6, NPI-Q-D, SWLS, and CES-D-10 for caregivers of Latinos with dementia are psychometrically sound for use in clinical practice.
- The English and Spanish caregiver-administered versions of the NPI-Q-D, QOL-AD, and GDS for Latino care recipients with dementia are psychometrically sound for use in clinical practice.
- Researchers may testing these scales’ measurement invariance and factorial structure among diverse Latinos in the future.

## Data Availability

All data produced in the present study are available upon reasonable request to the authors

## Acknowledgements

Dr. Perales-Puchalt thanks the national and local organizations that have partnered with him to conduct present and past research since 2015. The research team thanks research participants included in all stages of this research as well as anyone who has contributed directly and indirectly to this research. We also thank Visión y Compromiso, the UsAgainstAlzheimer’s A-List and the Alzheimer’s Association’s TrialMatch for sharing the opportunity to participate with the people they serve. The ideas and opinions expressed herein are those of the authors alone, and endorsement by the authors’ institutions or the funding agency is not intended and should not be inferred.

## Compliance with Ethical Standards

### Ethical approval

All study procedures were approved by the Institutional Review Board of the University of Kansas Medical Center (STUDY00145615).

### Informed consent

All caregiver participants gave written informed consent, as did those care recipients who were determined to be able to respond on their own via questions on comprehension of the informed consent.

